# Epidemiological Analysis of Lower Limb Vascular Trauma over 16 years in Brazil - A Nationwide View

**DOI:** 10.1101/2024.09.02.24312947

**Authors:** Carolina Carvalho Jansen Sorbello, Marcella Moura Ceratti, Felipe Soares Oliveira Portela, Marcelo Fiorelli Alexandrino da Silva, Marcelo Passos Teivelis, Antonio Eduardo Zerati, Nelson Wolosker

## Abstract

**Background:** Lower limb vascular trauma (LLVT) represents a significant public health challenge due to its potential to cause complex injuries that are difficult to manage, leading to increased morbidity, mortality and healthcare costs.

**Objective:** to investigate the incidence, lethality, population characteristics, and economic burden of LLVT in Brazil, the largest country in South America, from 2008 to 2023.

**Methods:** We used data from DATASUS (Department of Information and Informatics of the Brazilian Public Health System), which is the world’s largest public health system database. Our analysis focused on LLVT cases surgically treated in Brazil from 2008 to 2023. The study focused on demographic distribution, sex proportion, age groups, regional variations, hospital stays, intensive care unit (ICU) stays, lethality rates and financial expenditures.

**Results:** The study encompassed 20,349 LLVT cases and found a decrease in the number of cases over the years. LLVT was predominantly seen in males (70%), with an average patient age of 39.68 years. The Northeast and North regions registered the highest incidence, while the Southeast had the lowest. Most patients had a short hospital stay, averaging two days. The majority of patients did not need to be admitted to the ICU, and those who did stayed for an average of 4.48 days. The lethality was 5.96%, with bilateral LLVT showing a slightly lower mortality rate than unilateral cases. The total expenditure over 16 years, inferred by the amount passed on to SUS, totaled 9,537,664 USD, indicating a substantial economic impact.

**Conclusion:** LLVT has a significant impact on public health, mainly because it affects the economically active population, with a high risk of death or mutilating sequelae. Although there has been a general decrease in incidence, the persistence of high costs and high lethality rates indicate the need for targeted preventive measures. Future studies must investigate the causes and potential improvements in managing LLVT in Brazil.

## INTRODUCTION

Limb trauma is a major public health concern due to its significant impact on the health of those affected, which can lead to social, professional, and psychological damage. Much of the initial understanding of limb injuries comes from the time of the Great Wars. During this period, up to 50% of trauma cases (1,2) involved limb injuries, with the majority caused by penetrating wounds (81% from explosions and 17% from firearm injuries) (1).

In civilian populations, extremity injuries are mainly caused by falls (which account for 50-60% of lower limb injuries), industrial/professional accidents, and road crashes (3).

The worldwide literature widely agrees that vascular traumas are more prevalent in young males (4). Vascular injuries represented about 2% of all trauma cases (5), with LLVT corresponding to 18.5% of vascular traumas (4,5). The LLVT lethality is up to 29% (6). On average, patients with LLVT spent 6 days in the ICU, while those with upper limb vascular traumas spent 4.5 days (4,5). No population-based studies have determined the healthcare costs associated with lower limb vascular traumas.

It is important to note that this information is from studies in developed countries in the northern hemisphere, so it can not be extrapolated to developing or underdeveloped countries, let alone the global population. This is particularly true in less developed countries, where traffic accidents and violence rates are generally higher (7,8).

Brazil, the largest country in Latin America, is a developing nation and has just over 200 million inhabitants (9). The Brazilian health system (SUS - ‘Sistema ^Ú^nico de Saúde’) is considered the largest in the world, as anyone in the Brazilian territory has the right to be treated in this system. Recent statistics indicate that 71% of Brazilians rely exclusively on the SUS (10). All the data collected by the SUS, such as procedures, diagnoses, hospitalizations, outcomes, and demographic characteristics, is publicly available on DATASUS (Departamento de Informação e Informática do SUS), an online portal (11).

This study aimed to analyze data from the DATASUS on the incidence, lethality, demography, costs, and hospitalization time of LLVT in Brazil to present the context of LLVT in a non-developed country.

## METHODOLOGY

### Study Period, Data Source, and Extraction

Data was collected from DATASUS (11). The analysis focused on vascular trauma procedures of the lower limbs performed from 2008 to 2023. The total population analyzed in this study does not precisely correspond to the entire population of Brazil since some individuals use private health services, which are not accounted for in DATASUS. However, it can be inferred that the study’s total sample is greater than 140 million, corresponding to those who exclusively rely on the public health system (SUS) (10). The statistical calculations considered a population of 140 million individuals.

### Automated Data Extraction Process

The data collection process was automated utilizing a Python-based protocol (version 2.7.13; Beaverton, OR, USA) developed by our IT department. The extraction was performed on a Windows 10 system, using Selenium WebDriver (version 3.1.8; Selenium HQ) and Pandas (version 2.7.13; Lambda Foundry, Inc. and PyData Development Team, NY, USA) for data segregation and refining on the DATASUS platform.

### Procedure Selection on the Platform

The data on trauma was collected using ICD-10 codes (S00-S99 and T00-T98). It was then filtered by specific procedures: ‘Surgical treatment of bilateral traumatic vascular injuries of the lower limbs’ and ‘Surgical treatment of unilateral traumatic vascular injuries of the lower limbs’, associated with the following codes: 0406020442, 0406020450, 0406020507 and 0406020515. This filtering allowed the focus only on records related to lower limb vascular injuries caused by trauma. Neither amputations nor medically treated LLVT were included in this analysis.

The extracted information included the annual number of procedures, distribution by region, patient demographics (age and gender), length of hospitalization, ICU days, lethality and financial reimbursements. Financial data in Brazilian reais (R^$^) was converted to US dollars (U^$^) using the median exchange rate from 2008 to 2023, which was U^$^1 = R^$^3.2018 (12).

### Data Compilation and Statistical Analysis

The data was compiled and organized in .csv format using Microsoft Office Excel 2019 (Redmond, WA, USA). The Brazilian Institute of Geography and Statistics (IBGE) obtained population data by age group (13). Statistical analyses were conducted using SPSS version 20.0 for Windows (IBM Corp, Armonk, NY). Linear regression assessed trends in upper limb vascular trauma across age groups. Chi-square and likelihood ratio tests were employed to evaluate regional variations in trauma incidents and lethality. A p-value of ≤ 0.05 was set as the threshold for statistical significance.

### Ethical Approval

All data is anonymous and publicly available on the DATASUS online platform. Therefore, no informed consent from participants was necessary. This research received approval from the institution’s Research Ethics Committee (CAAAE 35826320.2.0000.0071).

## RESULTS

A total of 20,349 cases of LLVT occurred in Brazil between 2008 and 2023 (16 years), affecting approximately 90 out of every 100,000 people per year. Of the cases, 18,433 (90.6%) were unilateral, and 1,916 (9.4%) affected both lower limbs. The incidence of LLVT decreased from 2008 to 2023. In unilateral LLVT, there was an average decrease of 2.6% per year (mean decrease per year = 0.974; [0.970, 0.979]; p-value < 0.001), and in bilateral cases, the average decrease was 4.7% per year (ratio of average decrease = 0.953; [0.934, 0.973]; p-value < 0.001).

The incidence of LLVT over the years of the study can be observed in **Figure 1**.

**Figure 1:**
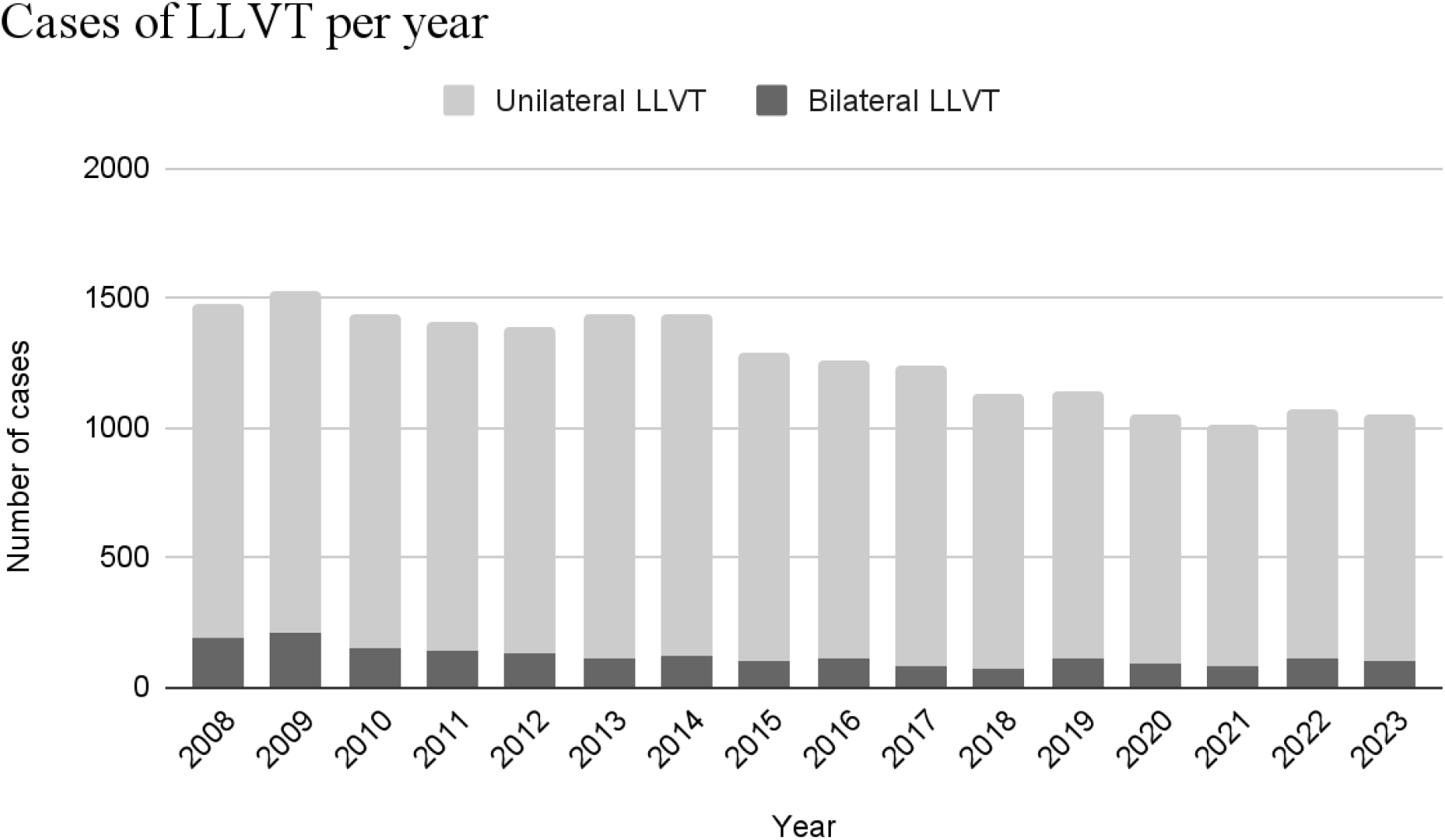
Cases of Lower Limb Vascular Trauma per year in Brazil. LLVT: Lower Limb Vascular Trauma.

**Table 1** shows the values and percentage of unilateral and bilateral LLVT in the 5 regions of Brazil. It is observed that most of the LLVT occurred in the Northeast and Southeast regions. However, when comparing with population size, the highest proportion of cases was observed in the Northeast and North, with approximately 13 cases per 100,000 inhabitants, followed by the South with 12 cases per 100,000 inhabitants, and the Midwest with 10 cases per 100,000 inhabitants. The region with the lowest number of cases per population was the Southeast, with 7 cases per 100,000 inhabitants over 16 years.

**Table 1:** Number of LLVT per region of Brazil. LLVT: Lower Limb Vascular Trauma.

| Region | Bilateral LLVT | Unilateral LLVT | p value |
| --- | --- | --- | --- |
| Midwest | 165 (8.6) | 1484 (8.1) | <0.001 |
| Northeast | 632 (33.0) | 6648 (36.1) |  |
| North | 315 (16.4) | 1881 (10.2) |  |
| Southeast | 542 (28.3) | 5207 (28.2) |  |
| South | 262 (13.7) | 3213 (17.4) |  |
| Total | 1916 (100.0) | 18433 (100.0) |  |

Males accounted for the majority of LLVT cases (75.4%): 73.6% of bilateral DVT cases and 75.6% of unilateral cases.

**Figure 2** shows the distribution of unilateral and bilateral LLVT cases by age group. There is a peak in incidence between the ages of 20 and 24, followed by a gradual decline until approximately 45 years of age, remaining relatively stable until 70 years. The age group with the least LLVT occurrence is those under 14, followed by those over 65 years for unilateral and bilateral LLVT. The mean age of patients with LLVT was 39.68 years. Furthermore, 13.7% of bilateral LLVT cases occurred in the 0 to 14 age group, whereas only 4.1% of unilateral LLVT cases occurred in this age group (p-value < 0.001). However, in the other age groups, the proportion of unilateral LLVT cases was greater than bilateral cases.

**Figure 2:**
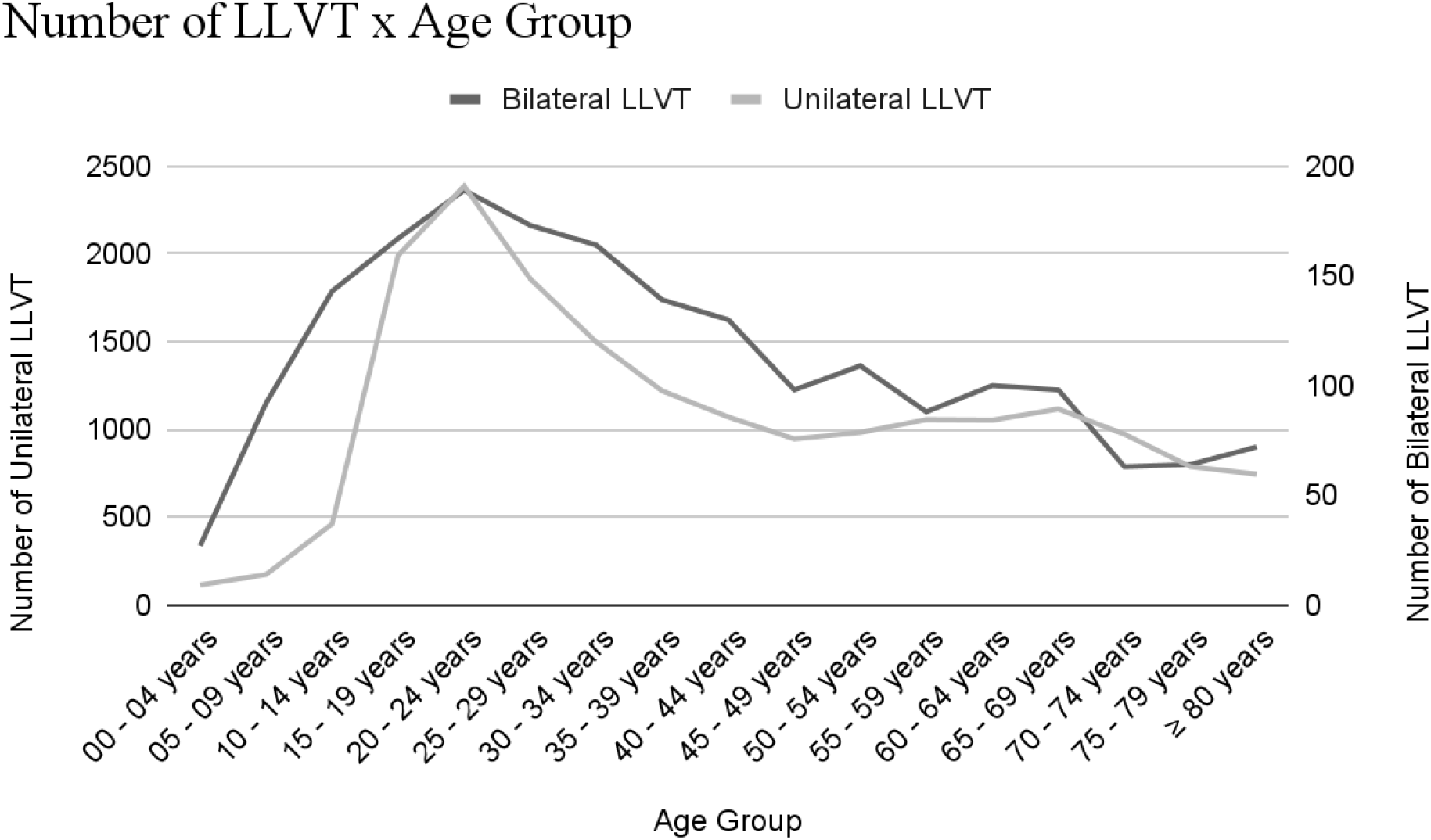
Cases of Lower Limb Vascular Trauma per Age Group. LLVT: Lower Limb Vascular Trauma.

**Figure 3** presents the number of hospitalization days for patients with LLVT. Most patients remained hospitalized for 2 days for both unilateral and bilateral LLVT.

**Figure 3:**
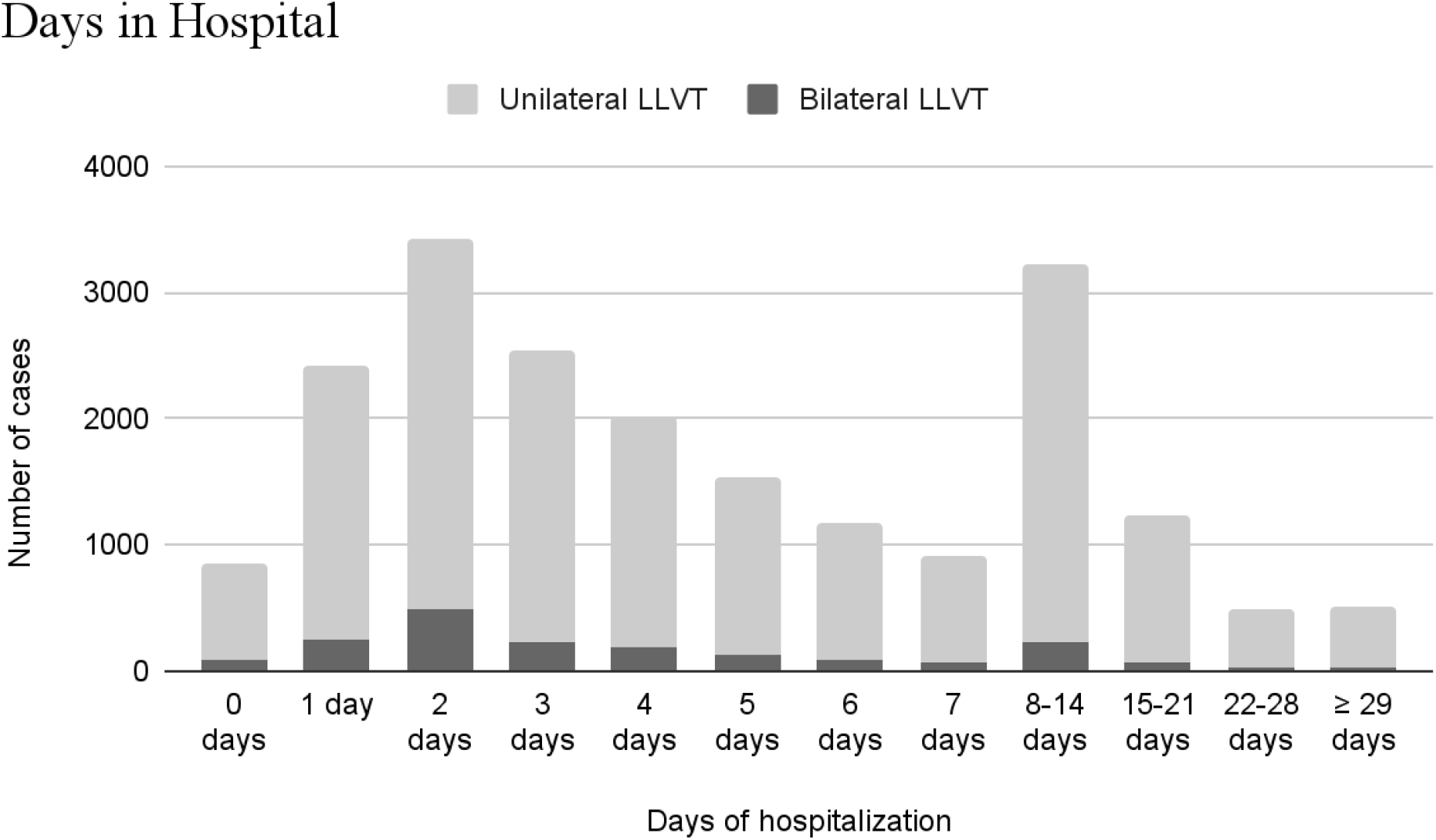
Days of hospitalization of patients with LLVT. LLVT: Lower Limb Vascular Trauma.

Patients with bilateral LLVT required fewer days of hospitalization compared to those with unilateral LLVT. The majority (43.4%) of patients with bilateral LLVT were hospitalized for 0 to 2 days, compared to only 30.8% of patients with unilateral LLVT (p-value < 0.001). In the unilateral LLVT group, the majority (40.6%) were hospitalized for 3 to 7 days. Only 7,0% of patients with bilateral LLVT, compared to 11.3% of those with unilateral LLVT, required hospitalization for 15 days or more (p-value < 0.001).

**Table 2** shows that most patients with LLVT did not require ICU admission (87,8% in bilateral LLVT and 82,3% in unilateral LLVT, p-value < 0.001). Among those requiring intensive care, the average number of days spent in the ICU was 4.48 days, among all patients with LLVT.

**Table 2:** Number of days in Intensive Care Unit (ICU) for patients with LLVT.

|  | LLVT |  | p-value |
| --- | --- | --- | --- |
|  | Bilateral | Unilateral |  |
| **Days in ICU** |  |  |  |
| 0 days | 1683 (87.8) | 15174 (82.3) | <0.001 |
| 1 to 5 | 185 (9.7) | 2529 (13.7) |  |
| 6 or more | 48 (2.5) | 730 (4.0) |  |
| Total | 1916 (100.0) | 18433 (100.0) |  |

**Table 3:** Cost of treatment for patients with LLVT from 2008 to 2023. LLVT: Lower Limb Vascular Trauma.

|  | **Total Cost**<br>(american dollars) | **Mean Cost per patient**<br>(american dollars) |
| --- | --- | --- |
| **Bilateral LLVT** |  |  |
| Region |  |  |
| Midwest | 61349.78 | 371.82 |
| Northeast | 208252.66 | 329.51 |
| North | 89283.21 | 283.44 |
| Southeast | 222673.25 | 410.84 |
| South | 124907.32 | 476.74 |
| **Total** | **706466.23** | **368.72** |
| **Unilateral LLVT** |  |  |
| Region |  |  |
| Midwest | 655403.39 | 441.65 |
| Northeast | 2973365.29 | 447.26 |
| North | 766978.29 | 407.75 |
| Southeast | 2668127.10 | 512.41 |
| South | 1767324.07 | 550.05 |
| **Total** | **8831198.14** | **479.10** |

Over the 16 years of the study, there were 1,212 deaths among patients with LLVT, corresponding to a lethality of 5.96%. The lethality of bilateral and unilateral LLVT were not statistically different, corresponding to 5.7% and 6%, respectively (p-value = 0.714).

**Figure 4** presents the lethality across the five regions of Brazil. The region with the highest lethality was the Southeast (7.31%), followed by the South (5.76%), Northeast (5.71%), Midwest (5.09%), and lastly, the North (4.19%), showing a statistically significant difference (p < 0.001). Interestingly, the lethality of bilateral LLVT was higher than unilateral LLVT in only two regions of Brazil, the Southeast and South.

**Figure 4:**
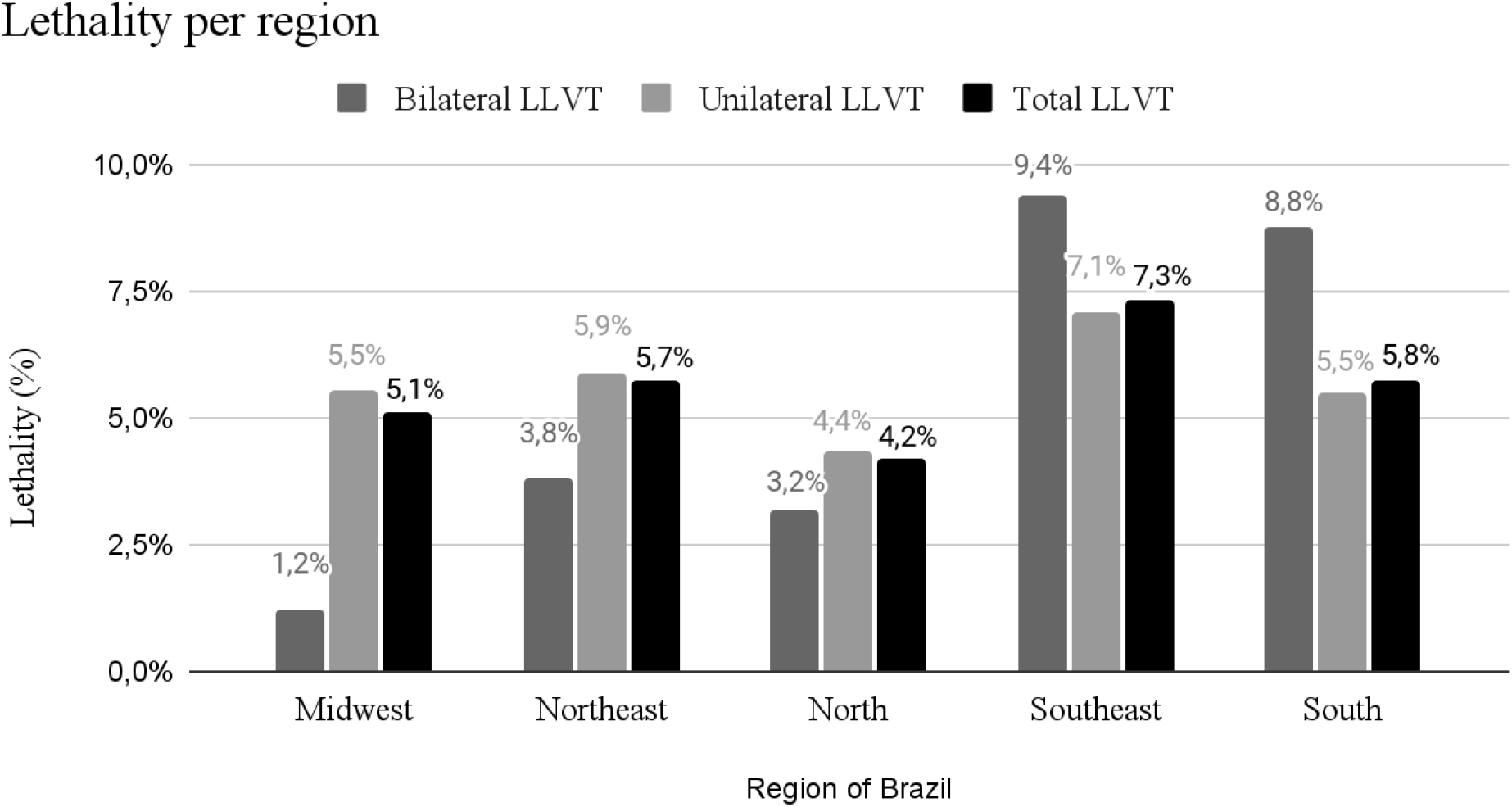
Lethality per region of Brazil. (p-value < 0.001). LLVT: Lower Limb Vascular Trauma.

The distribution of LLVT lethality by year is presented in **Figure 5**. The lethality of LLVT increased slightly over the years, with minor variations. In unilateral LLVT, there was an increase in lethality with a mean growth rate of 2.8% per year (lethality ratio of 1.028 per year [95% CI: 1.018, 1.039] and a p-value of <0.001). In bilateral LLVT, the increase in lethality was not statistically significant: lethality ratio of 1.021 per year [95% CI: 0.998, 1.044]; p-value = 0.080. Lethality for bilateral LLVT experienced greater fluctuations over the years, which can be explained by the smaller number of patients in this group.

**Figure 5:**
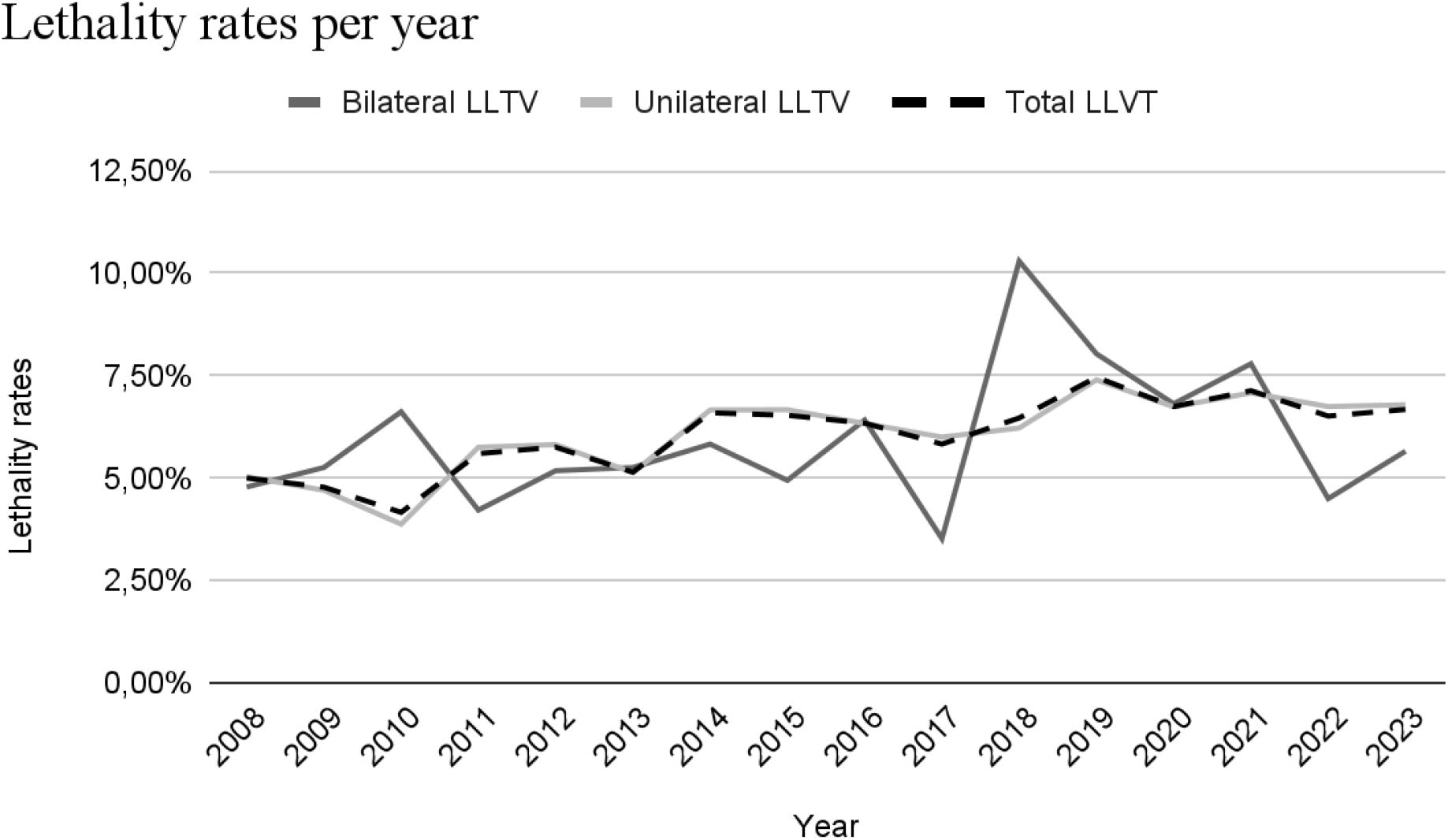
Lethality per year in Brazil. LLVT: Lower Limb Vascular Trauma.

The expenditure on acute LLVT treatment for the Brazilian public healthcare system from 2008 to 2023 was ^$^9,537,664 USD (dollars). The region that incurred the highest expense was the Northeast. **Table 4** shows the expenditures by region in Brazil, total and per patient. The total expenditure per patient with LLVT was ^$^469.70 USD. The region that spent the most per patient was the South (^$^545.53 USD per patient), while the region that spent the least was the Midwest (^$^390.99 USD per patient).

## DISCUSSION

Lower limb injuries can affect four main functional components: nerves, vessels, bones, and soft tissues. Vascular injury is particularly serious, as it can lead to hemorrhages, limb loss, death, and other disabling complications (4). Therefore, the management usually involves promptly addressing the injury in patients who show signs of vascular injury, such as pulsatile tumor, expanding hematoma, or distal ischemia (14).

The main causes of LLVT are firearm injuries, contusions caused by falls or traffic accidents, and stab wounds (15,16). In Brazil, the leading causes of LLVT are unknown. A study from Latin America analyzed just one center in Brazil and showed that the main cause of vascular trauma in all body regions was firearm injuries (17).

Recent studies have reported a higher prevalence of LLVT in Brazil than in other countries. For example, the United States reported 4,265 cases of LLVT over five years (4,5), while Brazil recorded 7,242 cases during the same frame despite having an approximately 50% smaller population (9,18). In comparison, a Scottish population study reported only 80 cases of LLVT over eight years (2011-2018) (6), while Brazil had 10,584 cases during the same period. Even after adjusting for population size, Brazil experienced approximately three times more LLVT cases than Scotland (19).

This study also observed a decline in the incidence of LLVT from 2011 to 2018. A plausible hypothesis for this trend is Brazil’s reduced traffic and occupational accidents over the past two decades (20,21). Additionally, the SARS-COV-19 pandemic may have contributed to this decrease, as the numbers in 2020 and 2021 were lower than in previous years (22).

In terms of Brazil’s macro-regions, the Northeast and North showed the highest incidence of LLVT per capita, while the Southeast had the lowest. This distribution may reflect a higher prevalence of accidents and physical violence in the North, Northeast, and Midwest compared to the Southeast and South (23,24).

Over 70% of LLVT cases in Brazil occurred in males, which is consistent with global findings, where approximately 70% to 80% of cases occur in males (5,25). This aligns with the higher rates of traffic accidents and violence among males (26).

The average age of LLVT patients in Brazil was 39.68 years, consistent with other global studies reporting an average age range of 31 to 50 years (27,28). This demographic is more prone to traffic accidents and violence (26). Accidents, violence, and chronic diseases are significant public health issues impacting a country’s economy (29).

The management of traumatic vascular injuries is well-documented in the literature and can be performed using endovascular or traditional approaches. For lower limb injuries, hemodynamically stable patients with less complex injuries can undergo endovascular treatment using angioplasty balloons and covered stents. Furthermore, angiotomography is the preferred method for diagnosing LLVT. Moreover, arteriography can also be used to support treatment planning and provide access to endovascular repair.

Conventional open treatment of arterial injuries can be categorized into primary amputation, vessel ligation, and revascularization. Primary amputation is typically used for damage control in unstable patients or cases of complex polytrauma with multiple injuries. Vessel ligation suits non-essential vessels, such as smaller veins and arteries. Primary arteriorrhaphy is performed if possible when revascularization is indicated, but in most cases, contralateral venous bypass grafts are required. A study of 74 patients with femoral artery trauma showed that only one of them was eligible for arteriorrhaphy, while bypass revascularization was necessary in 62 cases (30). Another alternative sometimes used is endovascular surgery (31,32).

Most LLVT patients in Brazil required two days of hospitalization. In contrast, the average hospital stay in the USA was 5.9 days for patients under 60 (4) and 13 days for older adults (5) In Australia, the average was 8 days (28). However, these figures are not directly comparable due to potential confounding injuries influencing the duration of hospital stays.

Interestingly, more patients with unilateral LLVT required intensive care treatment compared to those with bilateral LLVT (17.7% versus 12.2%, respectively). This discrepancy may be influenced by additional trauma-related injuries rather than just the lower limb vascular injury alone. The average ICU stay was 4.48 days in Brazil, compared to approximately 6 days in the USA (4,5).

It was initially hypothesized that bilateral LLVT would have higher lethality due to its association with high-energy trauma. Surprisingly, the lethality for bilateral LLVT was slightly lower than for unilateral, with 5.74% and 5.98%, respectively.

Lethality was higher in the Southeast and lower in the North, contradicting expectations of higher lethality in regions with less healthcare access and higher accident and violence rates, such as the North (23,24). LLVT lethality in Brazil was slightly lower than in the USA (6% versus approximately 8%) (4,5). In Scotland, lethality was significantly higher (approximately 21% of LLVT cases) (6), while in Australia it was substantially lower (0.3% of cases) (28).

Over the years, LLVT lethality in Brazil increased from 5% to approximately 6.7%. This trend may correlate with a dramatic rise in homicide rates driven by inequality, increased firearm availability, and drug use (24). Additionally, trend analysis of hospitalizations due to traffic accidents in São Paulo from 2000 to 2019 showed an initial rise in hospitalization rates, followed by stabilization and, in some cases, a resurgence, particularly among motorcyclists (33).

To date, no study has specifically evaluated the costs associated with LLVT. In Brazil, total expenditures of SUS amounted to ^$^9,537,664 over 16 years, representing a considerable cost per patient of ^$^469. The Southeast region spent the most on LLVT per patient, consistent with the greater availability of complex examinations and treatments in this region (34).

## LIMITATIONS

One of the limitations is the inability to define the exact population of the study. However, it is estimated to be greater than approximately 140 million, as this is the number of people who exclusively depend on SUS (10). Additionally, like any population-based database study, there is a risk of information bias since data relies on the records of healthcare professionals, which may result in underreporting of vascular traumas, especially if they are associated with other injuries.

In addition, the study did not consider LLVT managed with amputations or those managed clinically. However, it is known that the majority of LLVT are not managed clinically (35). Among those that are, they are rarely coded as “vascular trauma” in information systems based on clinical practice. Amputations are also a relatively uncommon method of managing LLVT (36). Consequently, the number of LLVT may have been slightly underestimated.

Finally, the costs assessed in this study may underestimate the total costs of treating LLVT since the reimbursement amount to SUS, defined by a fixed schedule, is based on what was presented in this research.

## CONCLUSION

Despite observed declines in the incidence of LLVT in recent years, concerning levels of lethality persist, especially in the Southeast region. More studies are needed to evaluate the causes of vascular traumas in Brazil so that better prevention strategies can be implemented.

## Data Availability

All data produced in the present study are available upon reasonable request to the authors

https://datasus.saude.gov.br/

## APPENDIX

**Supplementary Table 1:**
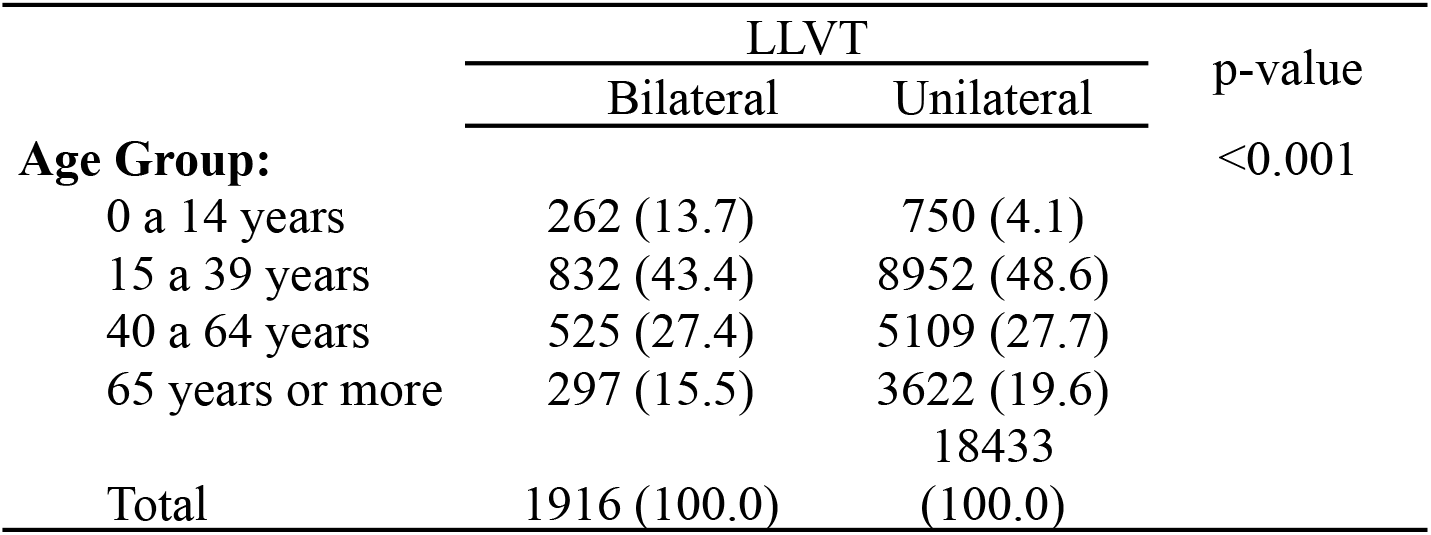
LLVT per age group.

**Supplementary Table 2:**
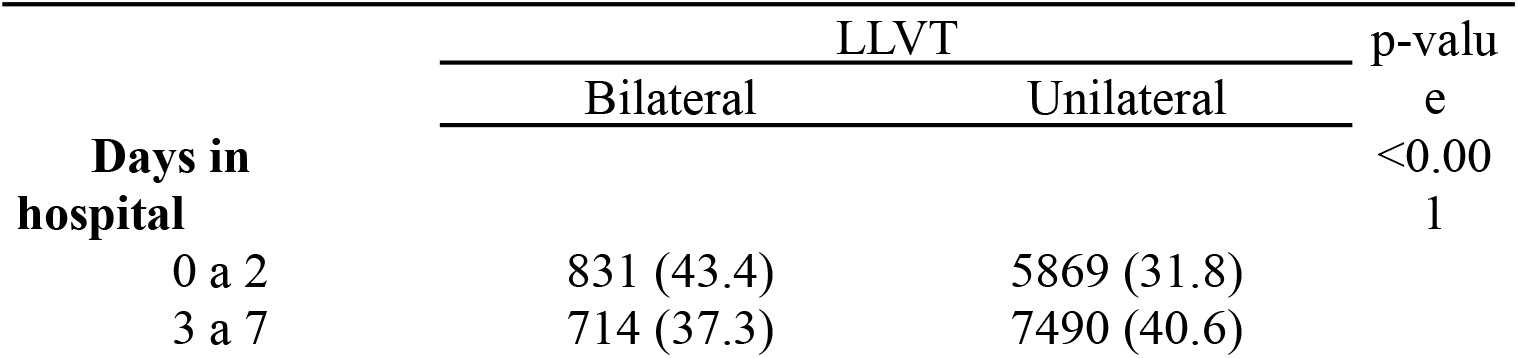
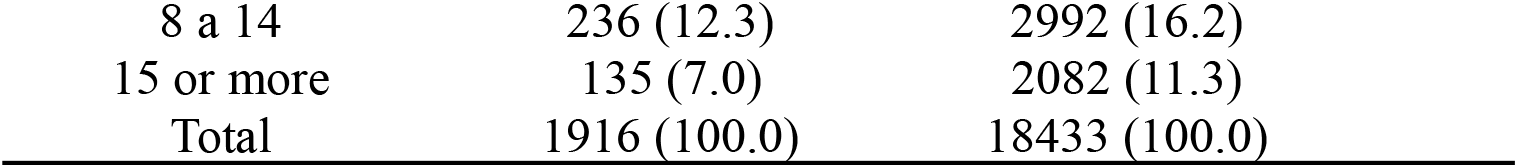
Days in hospital for patients with LLVT.

|  | LLVT |  | p-value |
| --- | --- | --- | --- |
|  | Bilateral | Unilateral |  |
| **Days in hospital** |  |  | <0.001 |
| 0 a 2 | 831 (43.4) | 5869 (31.8) |  |
| 3 a 7 | 714 (37.3) | 7490 (40.6) |  |

|  |  |  |
| --- | --- | --- |
| 8 a 14 | 236 (12.3) | 2992 (16.2) |
| 15 or more | 135 (7.0) | 2082 (11.3) |
| Total | 1916 (100.0) | 18433 (100.0) |

**Supplementary Table 3:** Lethality of LLVT per region of Brazil.

|  | Death |  | p-value |
| --- | --- | --- | --- |
|  | Yes | No |  |
| Bilateral LLVT |  |  |  |
| Midwest | 2 (1.2) | 163 (98.8) | <0.001 |
| Northeast | 24 (3.8) | 608 (96.2) |  |
| North | 10 (3.2) | 305 (96.8) |  |
| Southeast | 51 (9.4) | 491 (90.6) |  |
| South | 23 (8.8) | 239 (91.2) |  |
| Total | 110 (5.7) | 1806 (94.3) |  |
| Unilateral LLVT |  |  |  |
| Midwest | 82 (5.5) | 1402 (94.5) | <0.001 |
| Northeast | 392 (5.9) | 6256 (94.1) |  |
| North | 82 (4.4) | 1799 (95.6) |  |
| Southeast | 369 (7.1) | 4838 (92.9) |  |
| South | 177 (5.5) | 3036 (94.5) |  |
| Total | 1102 (6.0) | 17331 (94.0) |  |

## REFERENCES

1. Owens BD, Kragh JF, Wenke JC, Macaitis J, Wade CE, Holcomb JB. Combat wounds in operation Iraqi Freedom and operation Enduring Freedom. J Trauma. 2008 Feb;64(2):295–9.

2. Belmont PJ, Goodman GP, Zacchilli M, Posner M, Evans C, Owens BD. Incidence and epidemiology of combat injuries sustained during “the surge” portion of operation Iraqi Freedom by a U.S. Army brigade combat team. J Trauma. 2010 Jan;68(1):204–10.

3. Brasel KJ. Epidemiology. In: Moore EE, Feliciano DV, Mattox KL, editors. Trauma [Internet]. 8th ed. New York, NY: McGraw-Hill Education; 2017 [cited 2024 Aug 19]. Available from: https://accesssurgery.mhmedical.com/content.aspx?aid=1141184956

4. Barmparas G, Inaba K, Talving P, David JS, Lam L, Plurad D, et al. Pediatric vs adult vascular trauma: a National Trauma Databank review. J Pediatr Surg. 2010 Jul;45(7):1404–12.

5. Konstantinidis A, Inaba K, Dubose J, Barmparas G, Lam L, Plurad D, et al. Vascular trauma in geriatric patients: a national trauma databank review. J Trauma. 2011 Oct;71(4):909–16.

6. Thompson DC, Grossart C, Kerslake D, Tambyraja AL. Epidemiology and Outcomes of Peripheral and Non-Aortocaval Vascular Trauma in Scotland 2011 - 2018. Eur J Vasc Endovasc Surg Off J Eur Soc Vasc Surg. 2023 Mar;65(3):444–8.

7. Hazen A, Ehiri JE. Road traffic injuries: hidden epidemic in less developed countries. J Natl Med Assoc. 2006 Jan;98(1):73–82.

8. Bachani AM, Taber N, Mehmood A, Hung YW, Botchey I, Al-Kashmiri A, et al. Adolescent and Young Adult Injuries in Developing Economies: A Comparative Analysis from Oman and Kenya. Ann Glob Health. 2017;83(5–6):791–802.

9. Panorama do Censo 2022 [Internet]. [cited 2024 Jul 25]. Panorama do Censo 2022. Available from: https://censo2022.ibge.gov.br/panorama/

10. TabNet Win32 3.0: 47b - Estimativa da população SUS dependente (com base na Saúde Suplementar) [Internet]. [cited 2024 Aug 8]. Available from: https://tabnet.saude.sp.gov.br/tabcgi.exe?tabnet/ind47b_matriz.def

11. DATASUS – Ministério da Saúde [Internet]. [cited 2024 Aug 12]. Available from: https://datasus.saude.gov.br/

12. Receita Federal [Internet]. [cited 2024 Aug 8]. Conversão de dólares para reais. Available from: https://www.gov.br/receitafederal/pt-br/assuntos/meu-imposto-de-renda/tabelas/conversao

13. IBGE | Portal do IBGE | IBGE [Internet]. [cited 2024 Aug 19]. Available from: https://www.ibge.gov.br/

14. Hirsch AT, Haskal ZJ, Hertzer NR, Bakal CW, Creager MA, Halperin JL, et al. ACC/AHA 2005 guidelines for the management of patients with peripheral arterial disease (lower extremity, renal, mesenteric, and abdominal aortic): executive summary a collaborative report from the American Association for Vascular Surgery/Society for Vascular Surgery, Society for Cardiovascular Angiography and Interventions, Society for Vascular Medicine and Biology, Society of Interventional Radiology, and the ACC/AHA Task Force on Practice Guidelines (Writing Committee to Develop Guidelines for the Management of Patients With Peripheral Arterial Disease) endorsed by the American Association of Cardiovascular and Pulmonary Rehabilitation; National Heart, Lung, and Blood Institute; Society for Vascular Nursing; TransAtlantic Inter-Society Consensus; and Vascular Disease Foundation. J Am Coll Cardiol. 2006 Mar 21;47(6):1239–312.

15. Hafez HM, Woolgar J, Robbs JV. Lower extremity arterial injury: results of 550 cases and review of risk factors associated with limb loss. J Vasc Surg. 2001 Jun;33(6):1212–9.

16. Topal AE, Eren MN, Celik Y. Lower extremity arterial injuries over a six-year period: outcomes, risk factors, and management. Vasc Health Risk Manag. 2010 Dec 3;6:1103–10.

17. Sonneborn R, Andrade R, Bello F, Morales-Uribe CH, Razuk A, Soria A, et al. Vascular trauma in Latin America: a regional survey. Surg Clin North Am. 2002 Feb;82(1):189–94.

18. Bureau UC. Census.gov. [cited 2024 Aug 8]. American Community Survey (ACS). Available from: https://www.census.gov/programs-surveys/acs

19. Scotland’s Census [Internet]. [cited 2024 Aug 19]. Scotland’s Census 2022 - Rounded population estimates. Available from: https://www.scotlandscensus.gov.uk/2022-results/scotland-s-census-2022-rounded-populatio n-estimates/

20. Almeida FS e S de, Morrone LC, Ribeiro KB. [Trends in incidence and mortality due to occupational accidents in Brazil, 1998-2008]. Cad Saude Publica. 2014 Sep;30(9):1957–64.

21. Andrade FR de, Antunes JLF. Trends in the number of traffic accident victims on Brazil’s federal highways before and after the start of the Decade of Action for Road Safety. Cad Saude Publica. 2019 Aug 29;35(8):e00250218.

22. Steinman M, de Sousa JHB, Tustumi F, Wolosker N. The burden of the pandemic on the non-SARS-CoV-2 emergencies: A multicenter study. Am J Emerg Med. 2021 Apr;42:9–14.

23. Guimarães RA, de Sena KG, de Morais Neto OL, Malta DC. Magnitude and factors associated with motor road traffic injuries in Brazil: Results from the National Health Survey, 2019. Injury. 2023 Mar 11;S0020-1383(23)00244-9.

24. Murray J, Cerqueira DR de C, Kahn T. Crime and violence in Brazil: Systematic review of time trends, prevalence rates and risk factors. Aggress Violent Behav. 2013 Sep;18(5):471–83.

25. Thomson I, Muduioa G, Gray A. Vascular trauma in New Zealand: an 11-year review of NZVASC, the New Zealand Society of Vascular Surgeons’ audit database. N Z Med J. 2004 Sep 10;117(1201):U1048.

26. Sorenson SB. Gender disparities in injury mortality: consistent, persistent, and larger than you’d think. Am J Public Health. 2011 Dec;101 Suppl 1(Suppl 1):S353–358.

27. DuBose JJ, Savage SA, Fabian TC, Menaker J, Scalea T, Holcomb JB, et al. The American Association for the Surgery of Trauma PROspective Observational Vascular Injury Treatment (PROOVIT) registry: multicenter data on modern vascular injury diagnosis, management, and outcomes. J Trauma Acute Care Surg. 2015 Feb;78(2):215–22; discussion 222-223.

28. Friend J, Rao S, Sieunarine K, Woodroof P. Vascular trauma in Western Australia: a comparison of two study periods over 15 years. ANZ J Surg. 2016 Mar;86(3):173–8.

29. Takala J, Hämäläinen P, Sauni R, Nygård CH, Gagliardi D, Neupane S. Global-, regional- and country-level estimates of the work-related burden of diseases and accidents in 2019. Scand J Work Environ Health. 2024 Mar 1;50(2):73–82.

30. Wolosker N, Guadêncio A, Kuzniec S, Rosoky RA, Kalume C, Neves CA, et al. Surgical treatment of noniatrogenic trauma of the femoral arteries. Sao Paulo Med J Rev Paul Med. 1996;114(1):1079–82.

31. Wolosker N, Nakano L, Anacleto MMM, Puech-Leão P. Primary utilization of stents in angioplasty of superficial femoral artery. Vasc Endovascular Surg. 2003;37(4):271–7.

32. Puech-Leão P, Kauffman P, Wolosker N, Anacleto AM. Endovascular grafting of a popliteal aneurysm using the saphenous vein. J Endovasc Surg Off J Int Soc Endovasc Surg. 1998 Feb;5(1):64–70.

33. Conceição GM de S, Alencar GP, Latorre M do RD de O. [Time trend in hospitalizations from motor vehicle accidents in the city of São Paulo, Brazil, 2000-2019]. Cad Saude Publica. 2021;37(11):e00036320.

34. Martins LOM, Dos Reis MF, Chaoubah A, Rego G. Ethnic-Regional Differences in the Allocation of High Complexity Spending in Brazil: Time Analysis 2010-2019. Int J Environ Res Public Health. 2023 Feb 9;20(4):3006.

35. Franz RW, Shah KJ, Halaharvi D, Franz ET, Hartman JF, Wright ML. A 5-year review of management of lower extremity arterial injuries at an urban level I trauma center. J Vasc Surg. 2011 Jun;53(6):1604–10.

36. Urrechaga E, Jabori S, Kang N, Kenel-Pierre S, Lopez A, Rattan R, et al. Traumatic Lower Extremity Vascular Injuries and Limb Salvage in a Civilian Urban Trauma Center. Ann Vasc Surg. 2022 May;82:30–40.

